# Biallelic loss of function variants in *WBP4*, encoding a spliceosome protein, result in a variable neurodevelopmental delay syndrome

**DOI:** 10.1101/2023.06.19.23291425

**Authors:** Eden Engal, Kaisa Teele Oja, Reza Maroofian, Ophir Geminder, Thuy-Linh Le, Evyatar Mor, Naama Tzvi, Naama Elefant, Maha S. Zaki, Joseph G. Gleeson, Kai Muru, Sander Pajusalu, Monica H. Wojcik, Divya Pachat, Marwa Abd Elmaksoud, Won Chan Jeong, Hane Lee, Peter Bauer, Giovanni Zifarelli, Henry Houlden, Orly Elpeleg, Chris Gordon, Tamar Harel, Katrin Õunap, Maayan Salton, Hagar Mor-Shaked

## Abstract

Over two dozen spliceosome proteins are involved in human diseases, also referred to as spliceosomopathies. WBP4 (WW Domain Binding Protein 4) is part of the early spliceosomal complex, and was not described before in the context of human pathologies. Ascertained through GeneMatcher we identified eleven patients from eight families, with a severe neurodevelopmental syndrome with variable manifestations. Clinical manifestations included hypotonia, global developmental delay, severe intellectual disability, brain abnormalities, musculoskeletal and gastrointestinal abnormalities. Genetic analysis revealed overall five different homozygous loss-of-function variants in *WBP4*. Immunoblotting on fibroblasts from two affected individuals with different genetic variants demonstrated complete loss of protein, and RNA sequencing analysis uncovered shared abnormal splicing patterns, including enrichment for abnormalities of the nervous system and musculoskeletal system genes, suggesting that the overlapping differentially spliced genes are related to the common phenotypes of the probands. We conclude that biallelic variants in *WBP4* cause a spliceosomopathy. Further functional studies are called for better understanding of the mechanism of pathogenicity.

## INTRODUCTION

The spliceosome is a complex of RNA and proteins responsible for promoting accurate splicing. More than two dozen spliceosome proteins are known to be involved in human diseases, also referred as spliceosomopathies. Despite the fundamental function of the spliceosome in all tissues, spliceosomopathies are typically tissue specific, resulting in defects affecting only one cell type.^1^ For example, the widely expressed SNRNP200 is part of the U4/U6 complex, and is a known cause of Retinitis pigmentosa 33, which affects exclusively the retina.^2^ This observation suggests that the particular components of the spliceosome play a crucial role in accurately splicing specific targets, which can lead to the development of distinct pathologies.

WBP4 (WW Domain Binding Protein 4), previously known as FBP21, is a 376-amino acid spliceosome protein that contains a zinc finger motif and two tandem-WW domains. It was first detected as part of the early spliceosomal complex, where it interacts with the U2-associated protein SF3B4, SIPP1, and the core splicing protein SmB/B’. These interactions are facilitated by the transient multivalent recognition of the WW domains with proline-rich sequences.^3–6^ Later it was found to be present exclusively in the Spliceosome B complex (B complex-specific proteins) which acts directly before spliceosomal catalytic activation. WBP4 was found to bind and inhibit SNRNP200 similarly to C9ORF78 and TSSC4.^7, 8^ In the transition from the B to the Bact complex, the RNA helicase SNRNP200 unwinds the U4/U6 duplex, releasing the U4 snRNP, and enabling the U6 snRNA to base-pair with the U2 snRNA, forming a catalytically important stem–loop.^9, 10^ Thus, inhibition of SNRNP200 plays a crucial role in splice site selection, which positions WBP4 as a key player with a vital role in determining the outcome of splicing.

Overall, WBP4 was shown to enhance splicing in vitro and in vivo,^5^ and to regulate alternative splicing,^11^ but was not described before in the context of human pathologies. Here we report a severe neurodevelopmental syndrome with variable manifestations caused by homozygous loss of function (LOF) variants in the *WBP4* gene in eight unrelated families.

## MATERIAL AND METHODS

### Ethics statement

The study was conducted in accordance with IRB-approved protocol 0306-10-HMO, and in accordance with the Declaration of Helsinki and approved by the Research Ethics Committee of the University of Tartu (Certificate No 269/M-18 on 17.04.2017). Informed consent for participation and for publication of facial photographs was obtained from the patient’s parents.

### Exome sequencing

Duo exome sequencing was performed for Family 1. Exonic sequences from DNA were enriched with the SureSelect Human All Exon 50 Mb V5 Kit (Agilent Technologies, Santa Clara, California, USA) (proband F1-II:1), or xGen Exome Research Panel v2 kit (Integrated DNA Technologies, IDT) (proband F1-II:4). Sequences were generated on a NovaSeq6000 sequencing system (Illumina, San Diego, California, USA) as 150-bp paired-end runs. The FASTQs were uploaded into the Geneyx Analysis platform.^12^ Alignment and variant calling of single nucleotide variations (SNVs), structural variants (SVs), CNVs and repeats were called using Illumina DRAGEN Bio-IT, with hg19 as human reference genome. The resulting VCF files were comprehensively annotated on the Geneyx Analysis annotation engine, and presented for analysis, filtering and interpretation. Exome analysis of the probands yielded 40 and 120 million reads, with a mean coverage of 80 and 120X, in accordance.

For Family 2, trio exome sequencing was performed in clinical diagnostic settings at Tartu University Hospital. SureSelect Human All Exon V5 kit (Agilent Technologies, Santa Clara, California, USA) was used for exome enrichment of the genomic DNA extracted from blood. The enriched exome was sequenced on HiSeq 4000 (Illumina, San Diego, California, USA) platform. The data processing and variant calling pipeline followed Genome Analysis Toolkit’s best practice guidelines.^13^ The specifics of the in-house pipeline have been previously described.^14^ Exome sequencing of families 3-6 were sequenced and analyzed as published elsewhere.^15–17^

### Sanger sequencing

An amplicon containing the variant of interest was amplified by conventional PCR of genomic DNA, and analyzed by Sanger dideoxy nucleotide sequencing.

### Whole genome sequencing (Family 2)

Whole genome sequencing and data processing were performed by the Genomics Platform at the Broad Institute of MIT and Harvard. PCR-free preparation of sample DNA (350 ng input at >2 ng/ul) is accomplished using Illumina HiSeq X Ten v2 chemistry. Libraries are sequenced to a mean target coverage of >30x. Genome sequencing data was processed through a pipeline based on Picard, using base quality score recalibration and local realignment at known indels. The BWA aligner was used for mapping reads to the human genome build 38. Single Nucleotide Variants (SNVs) and insertions/deletions (indels) are jointly called across all samples using Genome Analysis Toolkit (GATK) HaplotypeCaller package version 3.4. Default filters were applied to SNV and indel calls using the GATK Variant Quality Score Recalibration (VQSR) approach. Annotation was performed using Variant Effect Predictor (VEP). Lastly, the variant call set was uploaded to seqr for collaborative analysis between the CMG and investigator. Loss of heterozygosity regions was calculated by Manta, Smoove and Delly programs.

### Diagnostic RNA sequencing

For Family 2, patient-derived fibroblasts were cultivated in AmnioMAX- c100. Human whole transcriptome sequencing was performed by the Genomics Platform at the Broad Institute of MIT and Harvard. The transcriptome product combines poly(A)-selection of mRNA transcripts with a strand-specific cDNA library preparation, with a mean insert size of 550bp. Libraries were sequenced on the HiSeq 2500 platform to a minimum depth of 50 million STAR-aligned reads. ERCC RNA controls are included for all samples, allowing additional control of variability between samples. STAR (version 2.5.3) aligner was used to map sequencing reads to GRCh38 reference genome; genes and transcripts were defined using GENCODE v26. To detect expression outliers STAR generated reads per gene counts of 40 unsolved pediatric patients with suspicion of a genetic disorder and 100 controls from GTEx were analyzed using R package OUTRIDER.^18^ Statistical significance was determined using a cut-off of 0.05 for the adjusted p-values. The results were analyzed collaboratively between Broad CMG and investigators from Tartu University Hospital.

### Cell lines

CCD-1092Sk cells (ATCC Number: CRL-2114) and primary fibroblasts cells from biopsy were grown in Dulbecco’s modified Eagle’s medium (DMEM) supplemented with 10% fetal bovine serum (FBS). Cells were maintained at 37°C and 5% CO2 atmosphere.

### Immunoblotting

For immunoblotting, cells were harvested and lysed with NP-40 1% lysis buffer, and 20 μg of the extracts were run on a 10% TGX Stain-free FastCast gel (BioRad) and transferred onto a nitrocellulose membrane. Antibodies used for immunoblotting were anti-WBP4 (Bethyl Laboratories # A305-693A-M), and anti-Actin.

### RNA Sequencing and analysis

RNA from primary fibroblasts was isolated using the GENEzol™ TriRNA Pure Kit (Geneaid). RNA samples for three biological replicates (approximately 1×10^6^ cells for family 1 and approximately 0.3×10^6^ cells for family 2) were subjected to sequencing. Raw reads were assessed using the FastQC tool v0.11.9. The processed reads were aligned to the human transcriptome and genome version GRCh38 with annotations from using STAR v2.7.10.^19^ Counts per gene quantification was done with featureCounts v2.0.1.^20^ Normalization and differential expression analysis were done with the DESeq2 package v 1.30.1.^21^ Pair-wise comparisons were tested with default parameters (Wald test), without applying the independent filtering algorithm. Significance threshold was taken as padj<0.05. In addition, significant DE genes were further filtered by the log2FoldChange value with lfcThreshold=1. rMATS v4.1.2^22^ was used to identify differential alternative splicing events. For each alternative splicing event, we used the calculation on both the reads mapped to the splice junctions and the reads mapped to the exon body (JCEC). Significance threshold was taken as FDR<0.05. In addition, significant splicing events were further filtered by the Inclusion level difference value for InclDiff > |0.1|. Gene-set enrichment analysis was performed using the GeneAnalytics tool.^23^

### Gene-phenotype relationship analysis

Gene-phenotype associations were calculated using Geneyx Analysis phenotyper tool.

## RESULTS

### Biallelic mutations in *WBP4* cause highly variable neurodevelopmental syndrome

To identify the genetic cause of a developmental delay and autism in a male child born to a consanguineous family (Figure 1A, F1-II:1), and of another affected offspring (individual F1-II:4; fetus with atrioventricular (AV)-canal, suspected absence of thymus, and intrauterine growth restriction), quad exome sequencing was performed. No pathogenic variants in genes known to be associated with neurological disorders were found, or any other known pathogenic events in both siblings. As the consanguineous background was suggestive for autosomal recessive inheritance, we focused on rare homozygous variants. A potentially harmful homozygous frameshift variant in *WBP4* was found in both siblings (chr13:g. 41072793 GA > G; NM_007187.5:c.499delA; p.(Thr167fs)(hg38)), predicted to result in a premature stop codon. Segregation study by Sanger sequencing revealed that the parents were heterozygous carriers, affected individuals were homozygous for the variant, and two unaffected siblings were either heterozygous or homozygous wild type (WT) (Figure 1A and Figure S1). This variant is absent from the Genome Aggregation Database (gnomAD), and no other homozygous LOF variant in this gene was described before.

**Figure 1.**
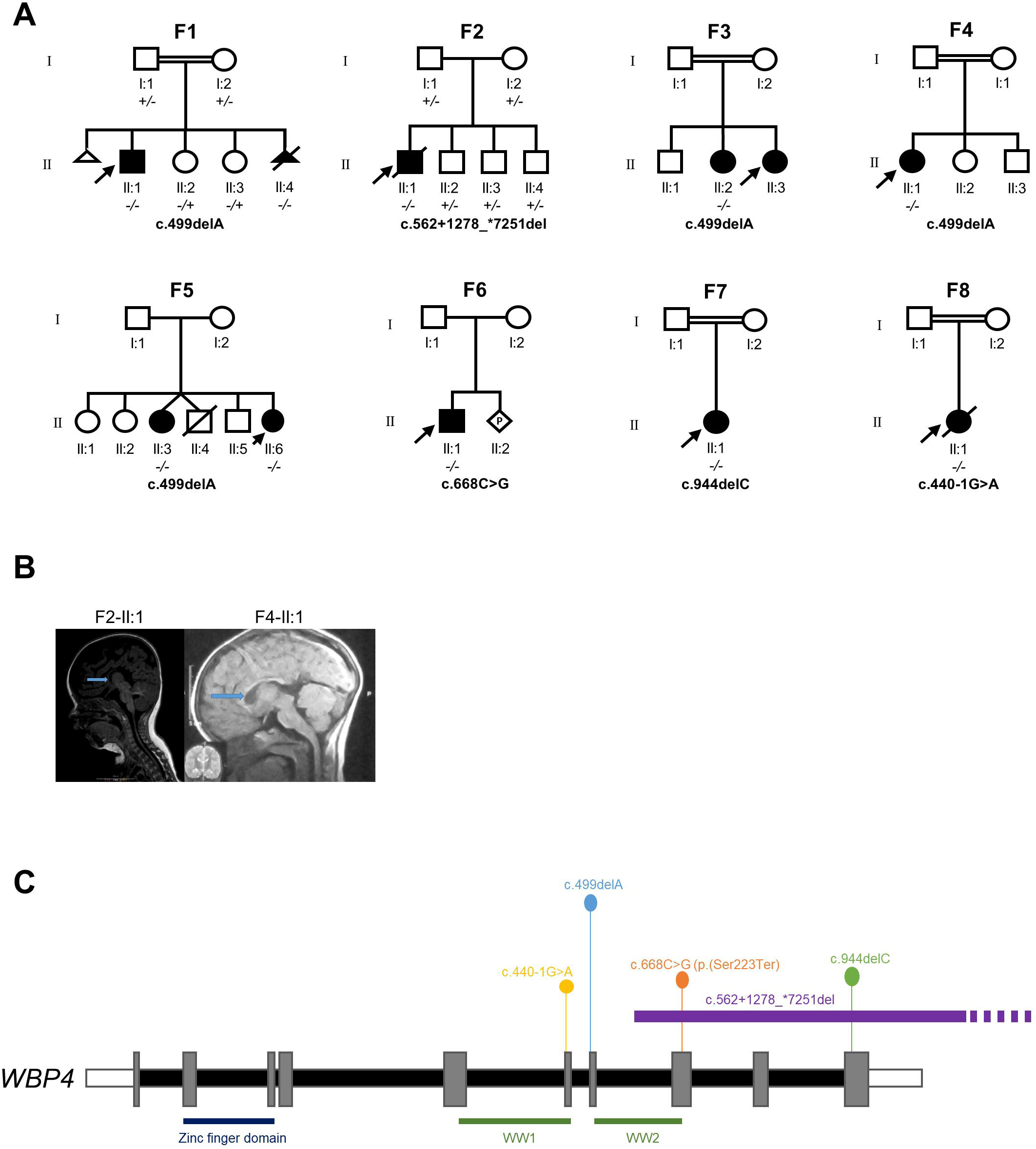
Clinical and genetic findings in eight families with *WBP4*-related syndrome. A. Pedigrees of eight families with *WBP4*-related syndrome. Symbols: filled black – affected individuals; diagonal line – deceased; double line – consanguinity; arrows indicate the probands. In each family the likely pathogenic variant in *WBP4* is written beneath the pedigree using NM_007187.5 transcript. Homozygotes for the likely pathogenic variant are noted as -/- and heterozygotes as +/-. F2:II-1 died of aspiration pneumonia in childhood. F5-II:4 died of intestinal obstruction in infancy; the parents of the proband (I:1 and I:2) are from the same village. F8-II:1 died in infancy due to cardiopathy. B. Magnetic resonance images (MRI) of two patients during their early childhood. MRI of F2-II:1 shows agenesis of corpus callosum (arrow) and a midsagittal T1-weighted MRI of F4-II:1 shows hypoplastic corpus callosum (arrow). C. Schematic diagram of *WBP4* transcript NM_007187.5 and the five LOF variants identified in our study. The gray rectangles represent exons 1-10 starting from left to right. The functional domains (zinc finger, WW1, WW2) of the protein are drawn below as blue and green rectangles.

Through GeneMatcher^24^, a total of 11 affected individuals from eight families were identified, all carrying homozygous LOF variants (Table 1 and Figure 1A, 1C). All variants were absent or had an extremely low frequency (<0.01%) in public databases.

**Table 1.** Summary of the demographic, genetic and clinical features of individuals with *WBP4*-related syndrome. All patients are homozygous for the variant in *WBP4* and the frequency of characteristics is presented as a ratio of patients with the feature to patients with data.

| Characteristic | Individuals with *WBP4*-related syndrome (n=11) |
| --- | --- |
| Age at last investigation, years (n=8) |  |
| Median | 3 |
| Range | 1.25 to 10 |
| Gender |  |
| Female | 8/11 |
| Ethnicity |  |
| Egyptian | 5/9 |
| Arab Muslim | 2/9 |
| Estonian | 1/9 |
| Indian | 1/9 |
| WBP4 variant (%) |  |
| c.499delA | 7/11 |
| c.562+1278_*7251del | 1/11 |
| c.668C>G | 1/11 |
| c.944delC | 1/11 |
| c.440-1G>A | 1/11 |
| Motor delay | 8/8 |
| Speech delay | 8/8 |
| Hypotonia | 7/7 |
| Dysmorphic facial features |  |
| Abnormality of the outer ear | 6/7 |
| Abnormal palate morphology | 4/8 |
| Hypertelorism | 4/6 |
| Fine hair | 3/7 |
| Feeding difficulties / Failure to thrive | 6/7 |
| Aplasia/Hypoplasia of the corpus callosum | 5/8 |
| Other morphological central nervous system abnormalities | 5/8 |
| Atypical behavior |  |
| Autistic features | 5/7 |
| Hyperactivity | 5/7 |
| Stereotypy | 4/7 |
| Sleep-wake cycle disturbance | 4/5 |
| Intellectual disability | 5/5 |
| Congenital heart defect | 4/7 |
| Prenatal issues | 3/10 |
| Urogenital abnormalities | 3/10 |
| Weight and height <-2 SD at last investigation | 3/6 |
| Skeletal abnormalities | 3/4 |

Interestingly, some of the patients had a much more severe phenotype. The patient from Family 2 was a male patient born as a first child from a complicated pregnancy to non-consanguineous parents. Bilateral hydronephrosis, intrauterine growth retardation (IUGR) and mild oligohydramnios were seen during pregnancy in ultrasound. The child was born prematurely at 35+5 weeks of gestation via emergency C-section. At birth, all his growth parameters were below the 3rd percentile (weight 1616 g, length 40 cm, head circumference 29 cm). He had multiple congenital anomalies – anal atresia with fistula into bladder, distal hypospadia and cryptorchidism, bilateral hydronephrosis, congenital heart defect - atrial septal defect (VCC-ASD), agenesis of corpus callosum (CCA) and dysmorphic features (dolichocephaly, proptosis, hypertelorism, depressed nasal bridge, microretrognathia). Clinically, the VACTERL association was diagnosed. He presented failure to thrive, bilateral neurosensory hearing loss, optic nerve atrophy, convergent strabismus, global developmental delay, hypotonia, dysphagia and frequent infections.

Electroencephalogram showed altered electrical activity without epileptiform abnormalities, and brain Magnetic resonance imaging (MRI) confirmed the CCA and frontotemporal widening of subarachnoidal space (Figure 1B). Trio exome analysis was performed, but no pathogenic variants could be associated with the patient’s phenotype. In early childhood, trio genome sequencing, RNA sequencing from fibroblast cell cultures and untargeted metabolomics analysis from serum were additionally performed. RNA sequencing revealed a very low expression of the *WBP4* gene (Figure S3), and reanalysis of genome data indicated a homozygous deletion in this region (Figure S4). Homozygous deletion of the three last exons of the *WBP4* gene was validated by Sanger sequencing. The deletion is located in a ∼6.2 Mb size loss of heterozygosity (LOH)-region (precise coordinates for deletion chr13:41,074,140-41,090,168 (hg38)).

The most common clinical features among the affected individuals in eight families were global developmental delay, muscular hypotonia, dysmorphic facial features and feeding difficulties or failure to thrive (Table 1). Morphological central nervous system abnormalities, atypical behavior and moderate to severe intellectual disability were present in almost half of the patients. In some cases congenital abnormalities of the heart, gastrointestinal, skeletal and/or urogenital system were evident. The individual clinical features are described in detail in supplementary materials (Table S1).

### RNA sequencing analysis

Considering the fact that WBP4 codes for a spliceosome protein, we wished to understand how splicing is altered in cells lacking WBP4. For this, we took a skin biopsy from affected individuals F1-II:1 and F2- II:1 and their parents. Primary fibroblasts were grown and protein was extracted to evaluate the amount of WBP4 in the probands relative to their heterozygote parents and WT fibroblasts. Our results show that in both cases the heterozygote parents show half of WBP4 protein amount relative to the WT control and that probands with homozygous *WBP4* mutations have total ablation of WBP4 protein (Figure 2A).

**Figure 2.**
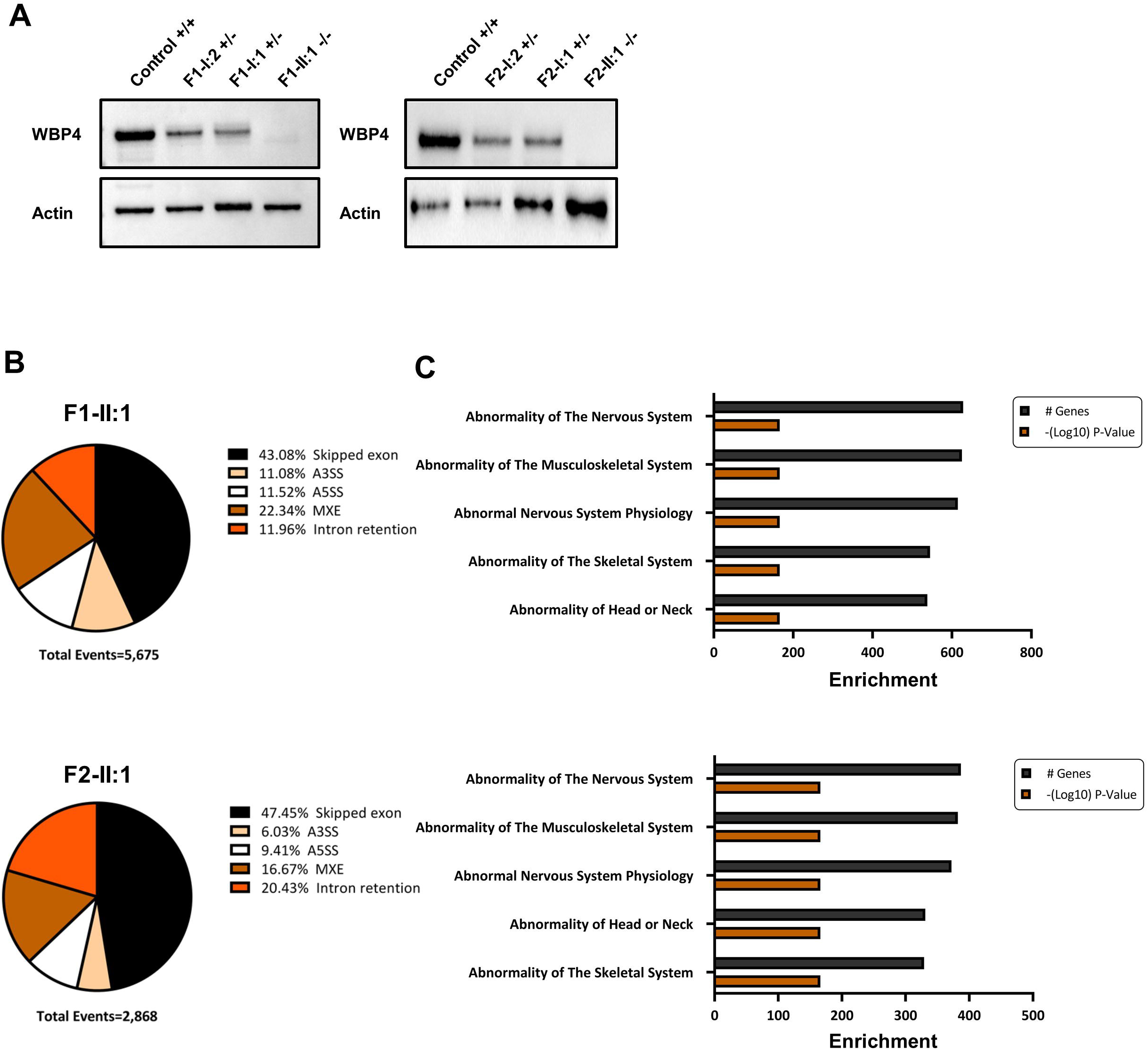
Immunoblotting and RNA sequencing analysis in Families 1 and 2. A. Western blot was performed to analyze the expression levels of WBP4 and actin proteins in fibroblast cell cultures from controls, family 1 (F1-I:2 mother, F1-I:1 father, F1-II:1 proband) and family 2 (F2-I:2 mother, F2-I:1 father, F2-II:1 proband). B. Summary of significant abnormal splicing events (FDR<0.05, IncLDiff < |0.1|) identified using rMATS in individual F1-II:1 (upper panel) and individual F2-II:1 (lower panel). C. Gene set enrichment for human phenotypes in individual F1-II:1 (upper panel) and individual F2-II:1 (lower panel). Enrichment is represented as the number of genes matching each human phenotype and the enrichment significance (-(Log10) P-value).

In addition, we extracted RNA from fibroblasts of the parents as well as probands and conducted RNA sequencing (RNA-seq). WBP4 mRNA amount was reduced in proband F1-II:1, as is expected by an early termination codon leading to nonsense mediated decay. In proband F2-II:1 expression was low and the transcript terminates at exon 7 out of 10 (NM_007187.5) as expected by the deletion location relative to the *WBP4* gene (Figure S5). Following analysis of the RNA-seq results we detected 579 genes with changes in gene expression in proband F1-II:1 relative to his parents (FDR<0.05, DEseq2^21^). To understand if these genes contribute to the proband’s phenotype, we used the GeneAnalytics tool^23^ to identify diseases related to the gene set. The human phenotype ontology pointed at enrichment for skin related phenotypes. Analyzing the results of the F2-II:1 proband we found 1,068 genes with altered gene expression as compared to his parents (FDR<0.05, DEseq2^21^) with a human phenotype enrichment of skin conditions as well as abnormalities in cardiovascular physiology (Figure S6A). We hypothesis that skin diseases enrichment which is not part of the probands’ phenotypes could suggest a difference in gene expression of skin-related genes between the young probands and their adult parents’ fibroblasts. Comparing the change in gene expression between the two probands we found 89 overlapping genes (P value<0.00001, Fisher exact test, Figure 2C) with no disease relevant gene set enrichment (Figure S6B and Table S2).

We have used rMATS (version 4.1.2) to identify abnormal splicing in the probands.^22^ Our analysis identified 5,675 differential splicing events in 2,859 genes in proband F1-II:1 compared to his parents (Figure 2B, upper panel, Table S3). All types of splicing events were present: skipped exons (2,445 events), alternative 5’ splice site (654 events), alternative 3’ splice site (629 events), mutually exclusive exons (1,268 events) and intron retention (679 events). To check if the differentially spliced genes are related to the proband’s phenotype, we used the GeneAnalytics tool^23^ and found enrichment for abnormalities of the musculoskeletal and the nervous system (Figure 2C, upper panel). Comparing proband F2-II:1 to his parents we identified 2,868 differential splicing events in 1,555 genes (Figure 2B, lower panel, Table S3). Again, all types of splicing events were identified: skipped exons (1,361 events), alternative 5’ splice site (270 events), alternative 3’ splice site (173 events), mutually exclusive exons (478 events) and intron retention (586 events). The human phenotype ontology pointed at genes related to the nervous system and the musculoskeletal system similar to proband F1-II:1 (Figure 2C, lower panel). While, WBP4 is expressed in all tissues it was found to be higher in the skeletal and heart muscles^25^ (Figure S8). This higher expression corresponds to the abnormal splicing of the musculoskeletal genes and to the heart anomaly - atrial septal defect in proband F2-II:1 (Table 1).

The principal components of the quantile normalized and standardized reads per exon show similarity between the two probands and the distinction of each proband from his parents (Figure 3A). Moreover, the probands samples are very distinct from samples of healthy, WBP4 +/+ children (Figure 3A). We continued to check the similarity in splicing events between the two probands and found 619 genes changing in splicing in both probands (Figure 3B, Figure S9A-C, and Table S3, P value<0.00001, Fisher exact test). To check if the common differentially spliced genes in both probands are related to their phenotype, we used the GeneAnalytics tool to perform human phenotype enrichment analysis. We found enrichment for abnormalities of the nervous system and musculoskeletal system, suggesting that the overlapping differentially spliced genes are related to the common phenotypes of the probands (Figure 3C). Among the differentially spliced genes related to the probands’ phenotype we identified for example *HUWE1*, *SMARCA2*, *SMARCC2*, *HNRNPH1* and *TCF4*. To understand if a connection exists between regulation of expression to that of splicing by WBP4 we compared the genes changing in expression to that of splicing. We could find a small but significant overlap that can be a result of an abnormal splicing event leading to a change in gene expression, as an inclusion of an exon or a retained intron harboring a stop codon (Figure S9D-F).

**Figure 3.**
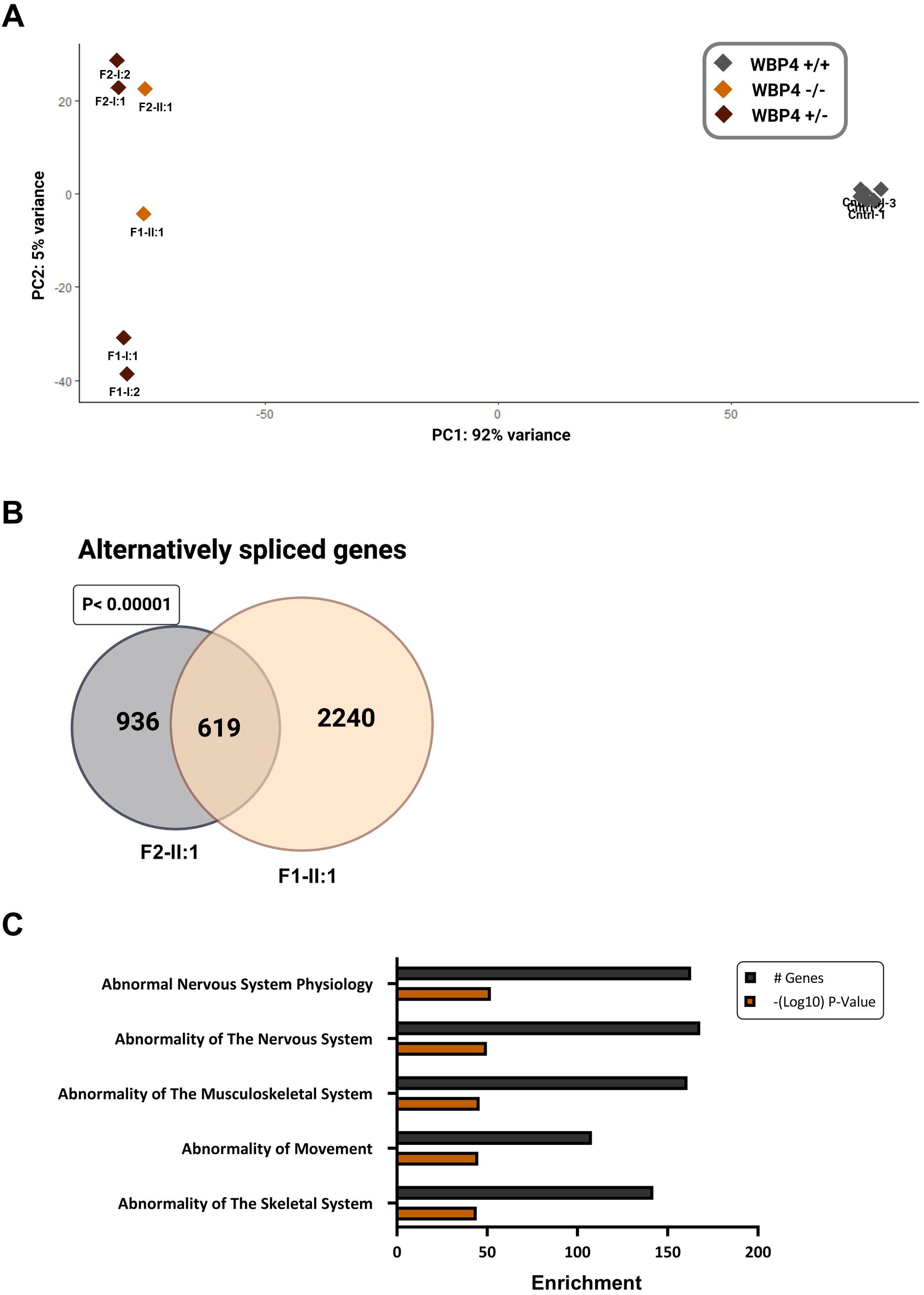
Common abnormal splicing events in F1-II:1 -/- and F2-II:1 -/-. A. RNA was extracted from fibroblasts of F1-I:2 +/-, F1-I:1 +/-, F1-II:1 -/-, F2-I:2 +/-, F2-I:1 +/- and F2-II:1 -/- fibroblasts and subjected to sequencing. Principal component analysis plot of PC1 and PC2 representing the variance between normalized exon read-counts in the two probands and their *WBP4* +/- relatives. B. Venn diagram representing the overlap in splicing between the two affected individuals (F1-II:1 and F2-II:1). C. Gene set enrichment analysis for 619 overlapping abnormally spliced genes. Gene set enrichment for human phenotypes, enrichment is represented as the number of genes matching each human phenotype and the enrichment significance (-(Log10) P-value).

Overall, the significant and shared alterations in splicing patterns in both patients, despite the district ethnical change, point at a similar cause. As suggested by the role of WBP4 in the inhibition of SNRNP200 and its exclusive presence in the B complex, where it acts directly before spliceosomal catalytic activation, depletion of WBP4 in both patients leads to a similar pattern of abnormal splicing and a shared pathomechanism of *WBP4* loss of function.

In order to determine the clinical relevance of differentially expressed (DE) and aberrantly spliced (AS) genes that the two probands (F1-II:1 and F2-II:1) had in common (89 and 619 genes respectively), we performed gene-phenotype relationship analysis. We analyzed the association between DE and AS genes symbols to 19 phenotypes found in at least 3 patients (Table 1). The search resulted in 46 (51.6%) and 356 (57.5%) directly related DE and AS genes (in accordance). 19.5% and 30% of the DE and AS related genes had at least 10 out of 19 phenotypic matches (in accordance). In a combined analysis of both DE and AS genes the top 10 highest ranking hits were all alternatively spliced – *NFIX*, *TRIO*, *ANKRD11*, *SZT2*, *HUWE1*, *PIGT*, *SMARCC2*, *PEX1*, *QRICH1*, *HRAS* (Table S4). This indicates that the alternatively spliced genes are more strongly connected to the patients’ phenotype.

## DISCUSSION

97% of human genes undergo pre-mRNA splicing, a process catalyzed by the spliceosome, a multi subunit complex. The spliceosome is comprised of small noncoding RNA molecules (U1, U2, U4, U5 and U6) as well as proteins.^26–29^ Among the spliceosome proteins, WBP4 is part of a group that plays a critical role in splice site decisions. These B-complex-specific proteins act directly before spliceosomal catalytic activation, fixing the specific splicing pattern of a substrate through irreversible loss of U4. In the transition from the B to the Bact complex, the RNA helicase SNRNP200 unwinds the U4/U6 duplex, releasing the U4 snRNP and enabling the U6 snRNA to base-pair with the U2 snRNA and form a catalytically important stem–loop. Therefore, WBP4 is a key player in spliceosomal activation.

Precise and fine-tuned gene expression is crucial for proper brain development. Neurodevelopment regulatory pathways are coordinated in space and time to produce a well-connected neuronal network.^30^ Over two dozen spliceosome proteins have been linked to human diseases, which are collectively referred to as spliceosomopathies. The tissue-specific nature of most spliceosome components suggests that they are not functionally equivalent. Studying spliceosomopathies can increase our understanding of the spliceosome and its functions.

Here we report a highly variable neurodevelopment syndrome caused by homozygous loss of function variants in *WBP4*, in eight different families.

The clinical presentation of the different affected individuals was highly variable. Individuals F1-II:1 and F2-II:1 presented the two sides of the spectrum, with one displaying a severe phenotype leading to multiple malformations and premature death (F2-II:1) and one only showing much less severe phenotype, with mainly intellectual disability and motor delay. Nevertheless, the two individuals have shared alterations in splicing patterns, stemming from WBP4 loss of function. While these phenotypic differences might be due to the nature of the genetic variants (large deletion in F2-II:1 vs. a frameshift variant in F1-II:1), they could also be a characteristic of spliceosomopathies, as previously shown in *CWC27* mutations, which lead to a spectrum of conditions including retinal degeneration, short stature, craniofacial abnormalities, brachydactyly, and neurological defects.^31^ This is also supported by the fact that both patients were shown to have complete loss of WBP4 protein.

Seven out of the top ten alternatively spliced genes with highest phenotype match, are known autosomal dominant genes. This fits with the assumption that splicing alteration are incomplete, mimicking a dominant effect.

One individual F2-II:1 presented the VACTERL association that comprises patients with at least three of the following characteristic features – vertebral defects, anal atresia, cardiac defects, tracheo- esophageal fistula, renal anomalies, and limb abnormalities. It has previously shown that heterozygous loss of WBP11 function cause a variety of overlapping congenital malformations, including cardiac, vertebral, tracheo-esophageal, renal and limb defects.^32^ *WBP11*, similarly to *WBP4*, encodes a component of the spliceosome with the ability to activate pre-messenger RNA splicing. *WBP11* heterozygous null mice are small and exhibit defects in axial skeleton, kidneys and esophagus. Our results support the conclusion of Martin et al 2020^32^ that loss-of function *WBP11* and *WBP4* variants should be considered as a possible cause of VACTERL association.

Larger cohort of affected individuals is necessary to recapitulate the entire spectrum of the *WBP4*- related disorder, and further functional studies are called for better understanding of the mechanism of pathogenicity.

## Data Availability

All analyzed data consists of patient's personal data and is stored according to regulations of the institutions. Pseudonymized data is available on request.
The ClinVar accession number for the DNA variants data are:
SCV003922044 WBP4 NM_007187.5 c.499delA
SCV003922045 WBP4 NC_000013.11 g.41074134_41090164del
SCV003922046 WBP4 NM_007187.5 c.668C>G
SCV003922047 WBP4 NM_007187.5 c.944delC
SCV003922048 WBP4 NM_007187.5 c.440-1G>A.

## ACKNOWLEDGEMENTS

The authors wish to thank the families for their participation in this study.

This work is supported by Estonian Research Council grants PRG471 and PSG774. The Broad Institute Center for Mendelian Genomics (UM1HG008900) is funded by the National Human Genome Research Institute with supplemental funding provided by the National Heart, Lung, and Blood Institute under the Trans-Omics for Precision Medicine (TOPMed) program and the National Eye Institute. MHW is supported by K23HD102589.

## AUTHOR CONTRIBUTIONS

H.M.-S., K.T.O., M.S., and E.E. designed the study and wrote the paper. T.H. and K.O. supervised the study, and contributed to writing the paper. H.M.-S. (F1) recognized this disease as a new clinical entity with the F1. E.E. has performed the comparative RNA sequencing analyses, and O.G. grew fibroblasts, extracted RNA and performed the western blot analyses, both under the supervision of M.S. N.E., M.T., and O.E. (F1) provided genetic consultation and evaluation. K.M. and K.O. (F2) provided genetic consultation and evaluation. S.P., M.H-W., K.O. and K.T.O. (F2) performed sequencing and analysis of trio genome and RNA for F2. R.M (F3-6) coordinated the local clinical study. M.Z. (F4, F5) J.G. (F5) … D.P. (F6) provided genetic consultation and evaluation. C.G and T-L.L. (F7-8) performed sequencing and analyses and coordinated the local clinical study.

## DECLARATION OF INTERESTS

HMS is an employee of Geneyx Genomics. Other authors declare no conflict of interest.

## DATA AVAILABILITY

The ClinVar accession number for the DNA variants data are:

SCV003922044 WBP4 NM_007187.5 c.499delA

SCV003922045 WBP4 NC_000013.11 g.41074134_41090164del

SCV003922046 WBP4 NM_007187.5 c.668C>G

SCV003922047 WBP4 NM_007187.5 c.944delC

SCV003922048 WBP4 NM_007187.5 c.440-1G>A.

All analyzed data consists of patient’s personal data and is stored according to regulations of the institutions. Pseudonymized data is available on request.

## Notes

### Author Declarations

IRB of Hadassah Medical Organization gave ethical approval for this work (protocol 0306-10-HMO). The Research Ethics Committee of the University of Tartu gave ethical approval for this work (Certificate No 269/M-18 on 17.04.2017).

