## Supplementary Table 1 for "Biallelic loss of function variants in *WBP4*, encoding a spliceosome protein, result in a variable neurodevelopmental delay syndrome"

**Table S1. The demographic, genetic and clinical features of individuals with WBP4-related syndrome in eight families.**

| Family | | Family 1 | | Family 2 | Family 3 | | Family 4 | Family 5 | | Family 6 | Family 7 | Family 8 |
|---|---|---|---|---|---|---|---|---|---|---|---|---|
| **Affected individual** | | **II:1** | **II:4** | **II:1** | **II:2** | **II:3** | **II:1** | **II:3** | **II:6** | **II:1** | **II:1** | **II:1** |
| Age at last investigation/history taking | | 6-10 | NA | 0-5 | 0-5 | NA | 6-10 | 6-10 | 0-5 | 0-5 | 0-5 | 0-5 |
| Gender | | Male | Female fetus | Male | Female | Female | Female | Female | Female | Male | Female | Female |
| WBP4 Variant (NM_007187.5) | | c.499delA p.(Thr167ProfsTer4) | c.499delA p.(Thr167ProfsTer4) | c.562+1278_*7251del | c.499delA p.(Thr167ProfsTer4) | c.499delA p.(Thr167ProfsTer4) | c.499delA p.(Thr167ProfsTer4) | c.499delA p.(Thr167ProfsTer4) | c.499delA p.(Thr167ProfsTer4) | c.668C>G p.(Ser223Ter) | c.944delC p.(Pro315GlnfsTer55) | c.440-1G>A |
| Zygosity | | Hom | Hom | Hom | Hom | Hom | Hom | Hom | Hom | Hom | Hom | Hom |
| Prenatal issues on fetal scan | | No | IUGR, AVC, suspected absence of thymus | IUGR, oligo-hydramnios, bilateral hydro-nephrosis | No | NA | Normal | Part of twins; twin died in infancy | Normal | Mild ventriculo-megaly, polyhydramnios | NA | NA |
| Growth at birth | Weight | 2526 g (-1.8 SD) | NR | 1616 g (< -3 SD) | appropriate for gestational age | NA | 3000 g (-0.5 SD) | 2600 g (-1.5 SD) | 3200 g (0 SD) | NA | NA | NA |
| | Length | NA | NR | 40 cm (< -3 SD) | NA | NA | 47cm (-1.1 SD) | 46cm (-1.7 SD) | 46cm (-1.7 SD) | NA | NA | NA |
| | OFC | NA | NR | 29 cm (-3 SD) | 35 cm (+0 SD) | NA | 33 cm (-1.2 SD) | 32.5cm (-1.6 SD) | 33 cm (-1.2 SD) | NA | NA | NA |
| Apgar index | | 9/10 | NR | 5/7/7 (neonatal respiratory distress) | NA | NA | 9/10 | 8/10 | 9/10 | NA (depressed at birth) | NA | NA |
| Growth at last investigation | Weight | 24 kg (0.3 SD) | NR | 7.8 kg (-2.5 SD) | 12 kg (-1.2 SD) | NA | 22 kg (-2.4 SD) | 20 kg (-2.3 SD) | 11.3 kg (-1.7 SD) | NA | NA | NA |
| | Length/ height | 118 cm (-0.7 SD) | NR | 69 cm (< -3 SD) | not reported | NA | 125 cm (-2.1 SD) | 118 cm (-2.3 SD) | 89 cm (-1.6 SD) | NA | NA | NA |
| | OFC | 50 cm (-1.3 SD) | NR | 42 cm (< -3 SD) | 46 cm (-1.75 SD) | NA | 51.5 cm (-0.3 SD) | 51.8 cm (0 SD) | 48 cm (-0.3 SD) | NA | NA | postnatal micro-cephaly |
| Craniofacial features | Hyper-telorism | NA | NR | Yes | No | NA | Yes | Yes | Yes | No | NA | NA |

| **Family** | | **Family 1** | | **Family 2** | **Family 3** | | **Family 4** | **Family 5** | | **Family 6** | **Family 7** | **Family 8** |
|---|---|---|---|---|---|---|---|---|---|---|---|---|
| **Affected individual** | | **II:1** | **II:4** | **II:1** | **II:2** | **II:3** | **II:1** | **II:3** | **II:6** | **II:1** | **II:1** | **II:1** |
| | Palpebral fissures (slant) | NA | NR | downward | No | NA | upward | No | mild upward | No | NA | NA |
| | Nasal bridge | NA | NR | depressed | No | NA | prominent | depressed | NA | No | NA | NA |
| | Mouth | No | NR | smooth philtrum, high arched palate | No | NA | No | short philtrum, high arched palate | No | short philtrum, tented upper lip, thick lips, high narrow palate, crowding of teeth | NA | cleft lip and palate |
| | Ears | abnormal ear morphology | NR | abnormal ear morphology | No | NA | protruding small ears | protruding low-set ears | protruding ears | anteverted and large ears | NA | NA |
| | Hair | No | NR | No | No | NA | thin hair | thin hair | thin hair | no | NA | NA |
| | Other | | | turricephaly, proptosis | No | NA | flat forehead, arched eyebrows, broad chin | | | asymmetric skull, prominent metopic ridge, bitemporal narrowing, bilateral epicanthal folds, overhanging columnella | cranio-synostosis | NA |
| Skeletal | | NA | NR | sandal gap | No | NA | NA | NA | NA | pectus deformity, pes planus | NA | bilateral club foot |
| Develop-mental milestones | Motor delay | Yes | NR | Yes | Yes | NA | Yes | Yes | Yes | Yes | Yes | NA |
| | Age at independent walking (details) | > 15m (walks, climbs, difficulty running) | NR | not yet | > 15m (ataxic gait, frequent falling) | NA | > 15m (walks well, jumps) | > 15m (walks well) | > 15m (wide-based gait) | > 15m (not yet walking; can take steps with support) | not yet | NA |
| | Speech delay | Yes | NR | Yes | Yes | NA | Yes | Yes | Yes | Yes | Yes | NA |
| | Age at first words (details) | not yet (syllables) | NR | not yet | > 1y (<10 meaningful words) | NA | > 1y (few double syllable words) | > 1y (few words) | > 1y (few letters) | not yet | not yet | NA |
| | Age of complete | not yet | NR | not yet | NA | NA | not yet | > 5y fairly controlled | not yet | NA | NA | NA |

| **Family** | | **Family 1** | | **Family 2** | **Family 3** | | **Family 4** | **Family 5** | | **Family 6** | **Family 7** | **Family 8** |
|---|---|---|---|---|---|---|---|---|---|---|---|---|
| **Affected individual** | | **II:1** | **II:4** | **II:1** | **II:2** | **II:3** | **II:1** | **II:3** | **II:6** | **II:1** | **II:1** | **II:1** |
| | ly toilet trained | | | | | | | | | | | |
| Intellectual disability (IQ) | | severe (NA) | NR | severe (NA) | NA | NA | severe (35) | moderate (40) | severe (35) | NA | NA | NA |
| Autistic features | | Yes | NR | No | No | NA | present (CARS: 38) | present (CARS: 35) | present (CARS: 36) | bruxism | NA | NA |
| Abnormal behaviors (e.g. aggression, hyperactivity, stereotypy, etc.) | | Hyperactivity | NR | Stereotypy | No | NA | Stereotypy, hyperactivity | Stereotypy, hyperactivity | Nervousness, hyper-activity, aggression | Hyperactivity | NA | NA |
| Sleep disturbances | | not reported | NR | Sleep-wake cycle disturbance | NA | NA | Sleep-wake cycle disturbance | Sleep-wake cycle disturbance | Sleep-wake cycle disturbance | NA | NA | NA |
| Seizures | Clinical seizures | No | NR | No | No | NA | apneic spells with tonic; onset in childhood; controlled on oxacarbazepine) | No | No | No | NA | NA |
| | EEG findings | Normal | NR | Diffuse non-epileptiform abnormal activity, diffuse abnormally slow rhythms | Normal | NA | Left parietal epileptgenic disharges | Normal | Normal | Normal | NA | NA |
| Hypotonia | | Yes | NR | Yes | Yes | NA | Yes | Hypotonia in early infancy | Yes | Yes | NA | NA |
| Cerebral imaging | | Normal | NR | CCA, frontotemporally widened subarachnoid space | Normal | NA | Hypoplastic thin corpus callosum, prominent cortical sulci | Hypoplastic thin corpus callosum | Hypoplastic thin corpus callosum, mild prominent cortical sulci | NA | CCA, abnormal cerebral cortical gyration | Abnormal cerebral cortical gyration |
| Feeding diffficuties and/or FTT | | Yes | NR | Yes (gastrostomy | Not reported | NA | Yes | Yes | Yes | Yes | NA | NA |

| Family | Family 1 | | Family 2 | Family 3 | | Family 4 | Family 5 | | Family 6 | Family 7 | Family 8 |
|---|---|---|---|---|---|---|---|---|---|---|---|
| **Affected individual** | **II:1** | **II:4** | **II:1** | **II:2** | **II:3** | **II:1** | **II:3** | **II:6** | **II:1** | **II:1** | **II:1** |
| | | | feeding, frequent vomiting) | | | | | | | | |
| Hearing loss | not reported | NR | Congenital sensorineural hearing impairment | Bilateral conductive hearing loss to mid and high frequencies | NA | No | No | No | NA | NA | NA |
| Strabismus | No | NR | Yes | NA | NA | Infrequent | No | No | NA | NA | NA |
| Congenital heart defect | No | AVC | ASD | NA | NA | Mitral regurgitation, small PDA | No | No | | NA | Multiple VSDs; cardiac coarctation |
| Other | - | - | Urogenital abnormalities (hydronephrosis, renal cyst, cryptorchidism, hypospadias, anal atresia with fistula); Frequent infections (bronchitis, otitis) | - | NA | - | - | - | - | Clitoris hypotrophy; Hirschsprung disease | Clitoris hypertrophy |

**Abbreviations: (*add*): ASD – atrial septal defect; AVC – atrioventricular canal defect; CARS – childhood autism rating scale; CCA – agenesis of corpus callosum, FTT – failure to thrive, Hom – homozygous; IUGR – intrauterine growth retardation; NA – not available; NR – not relevant; OFC – occipitofrontal circumference; PDA – patent ductus arteriosus; VSD – ventricular septal defect**
