## Supplementary Figures 1-9 for "Biallelic loss of function variants in *WBP4*, encoding a spliceosome protein, result in a variable neurodevelopmental delay syndrome"

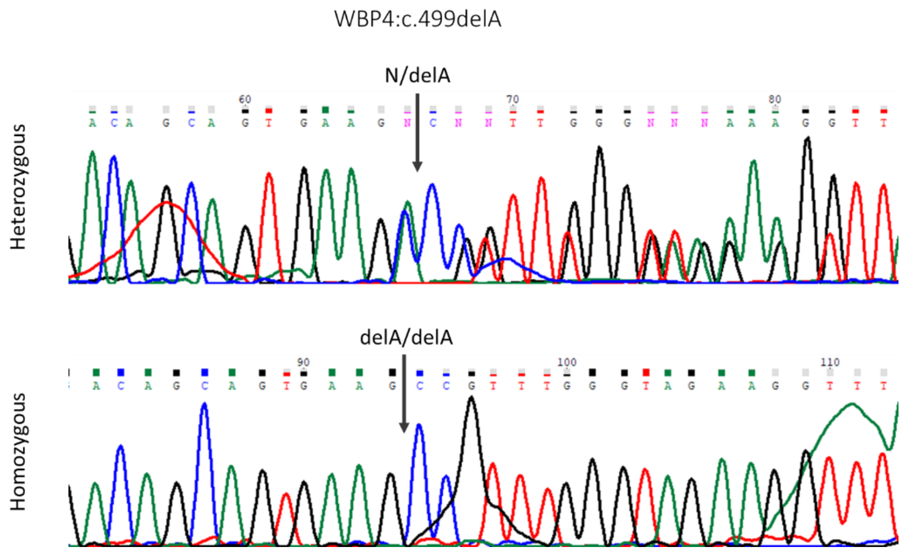


**Figure S1. Sanger sequencing of WBP4 c.499delA variant.** Heterozygous (upper panel) and homozygous (lower panel) individuals are F1-I:2 and F1-II:1 in accordance.

Figure S2. Haplotype analysis

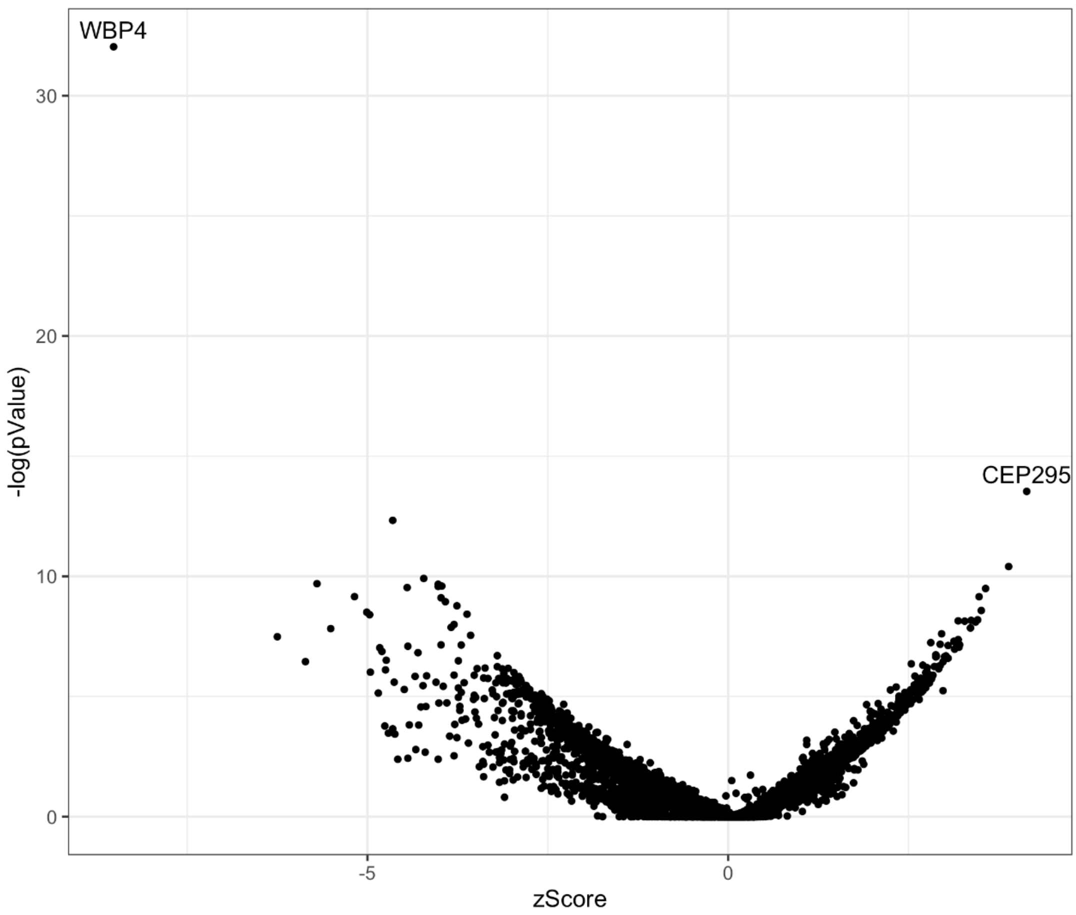


**Figure S3. Volcano plot of RNA sequencing results from fibroblast cell culture of F2-II:1** showing low expression of *WBP4* gene compared to 40 unsolved pediatric patients with suspicion of a genetic disorder and 100 controls from GTEx.

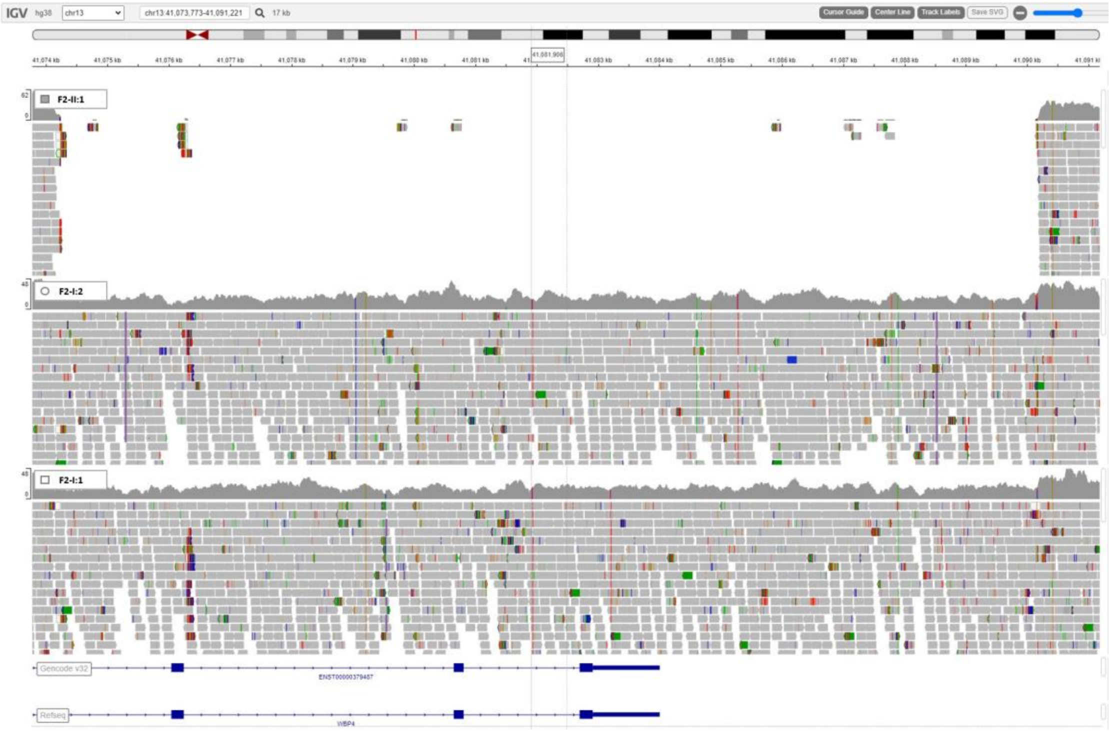


**Figure S4. Trio whole genome sequencing (WGS) in Family 2**. F2-II:1 in the first row has a homozygous deletion NC_000013.11:g.41074134_41090164del that covers the last three exons in the *WBP4* gene. The parents (F2-II:2 in second row and F2-II:1 in third row) have a heterozygous deletion.

**WBP4 Gene**

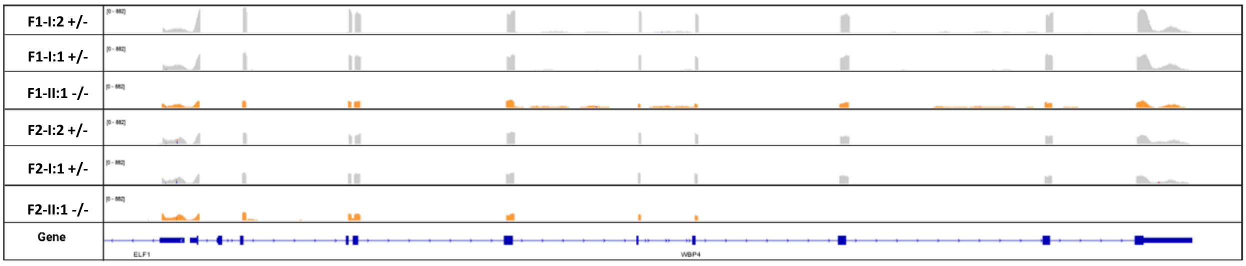


**Figure S5. Lower WBP4 transcript levels in F1-II:1 -/- and F2-II:1 -/-.**

RNA was extracted from F1-I:2 +/- , F1-I:1 +/-, F1-II:1 -/-, F2-I:2 +/-, F2-I:1 +/- and F2-II:1 -/- fibroblasts and subjected to sequencing. IGV browser snapshot of RNA-seq, peaks indicate relative expression levels at WBP4 gene.

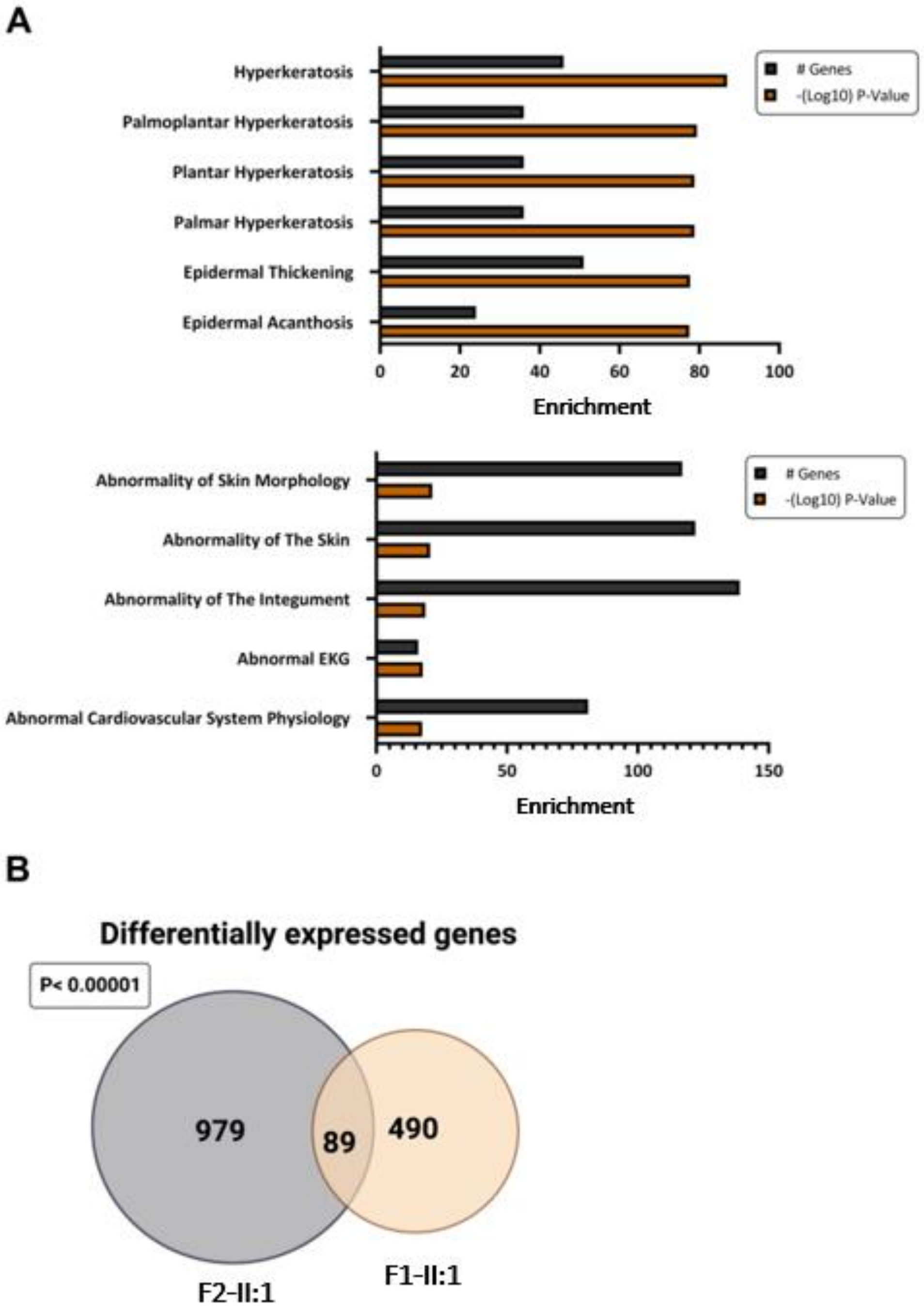


**Figure S6. Gene-expression profile in F1-II:1 -/- and F2-II:1 -/-.** A. RNA was extracted from two heterozygotes for the c.499delA variant (individuals F1-I:1 and F1-I:2, WBP4 +/-) and homozygote for the c.499delA variant (individual F1-II:1, WBP4 -/-), and subjected to sequencing. Differential expression analysis was performed followed by gene set enrichment for human phenotypes, enrichment is represented as the number of genes matching each human phenotype and the enrichment significance (-(Log10) P-value) (upper panel). RNA was extracted from two heterozygotes for the 16kbdel variant (individuals F2-I:1 and F2-I:2,

WBP4 +/-) and homozygote for the 16kbdel variant (individual F2-II:1, WBP4 -/-), and subjected to sequencing. Differential expression analysis was performed followed by gene set enrichment for human phenotypes, enrichment is represented as the number of genes matching each human phenotype and the enrichment significance (-(Log10) P-value). B. Venn diagram representing the overlap in expression between the two probands.

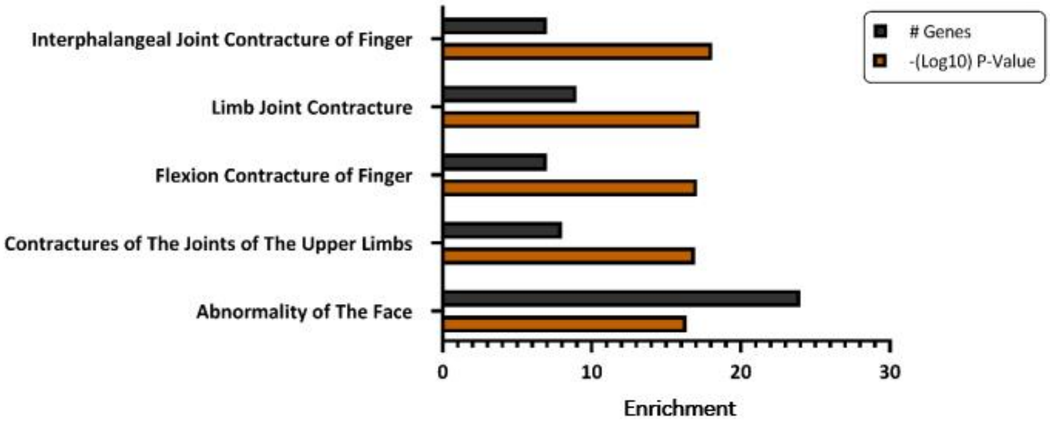


**Figure S7. Analysis of overlapping differentially expressed genes in** F1-II:1 -/- and F2-II:1 -/-. Gene set enrichment analysis for 89 overlapping differentially expressed genes. Gene set enrichment for human phenotypes, enrichment is represented as the number of genes matching each human phenotype and the enrichment significance (-(Log10) P-value).

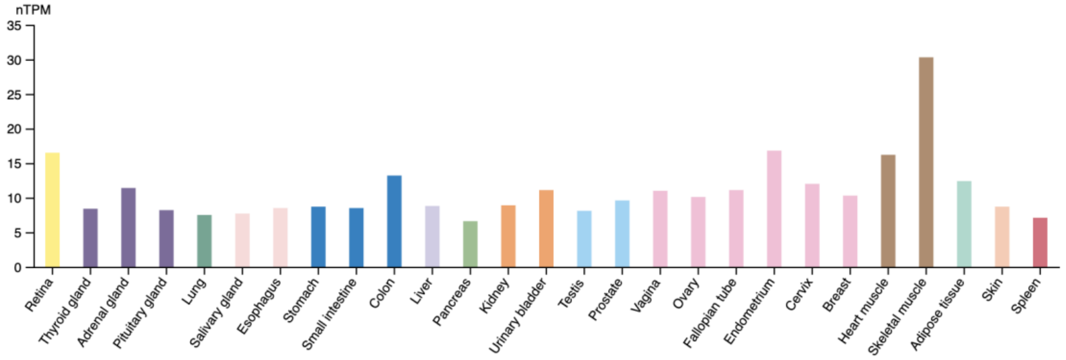


**Figure S8. WBP4 expression profile.** WBP4 mRNA levels per tissue from taken from the Genotype-Tissue Expression (GTEx) project.

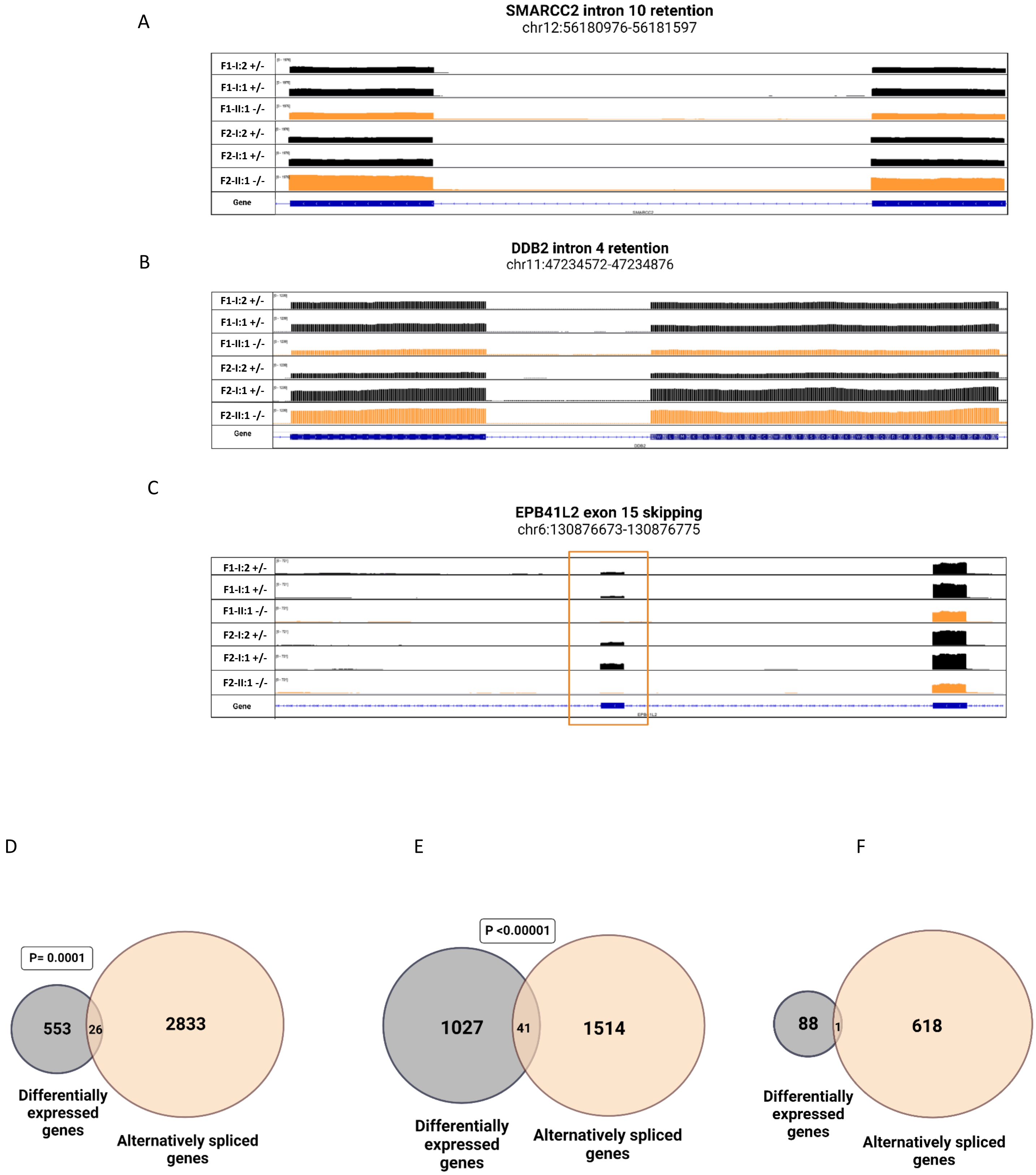
A
SMARCC2 intron 10 retention
chr12:56180976-56181597
F1-I:2 +/-
F1-I:1 +/-
F1-II:1 -/-
F2-I:2 +/-
F2-I:1 +/-
F2-II:1 -/-
Gene
B
DDB2 intron 4 retention
chr11:47234572-47234876
F1-I:2 +/-
F1-I:1 +/-
F1-II:1 -/-
F2-I:2 +/-
F2-I:1 +/-
F2-II:1 -/-
Gene
C
EPB41L2 exon 15 skipping
chr6:130876673-130876775
F1-I:2 +/-
F1-I:1 +/-
F1-II:1 -/-
F2-I:2 +/-
F2-I:1 +/-
F2-II:1 -/-
Gene
D
P= 0.0001
553
26
2833
Differentially expressed genes
Alternatively spliced genes
E
P <0.00001
1027
41
1514
Differentially expressed genes
Alternatively spliced genes
F
88
1
618
Differentially expressed genes
Alternatively spliced genes

**Figure S9. Splicing abnormalities in two affected individuals.** A-F: RNA was extracted from F1-I:2 +/- , F1-I:1 +/-, F1-II:1 -/-, F2-I:2 +/-, F2-I:1 +/- and F2-II:1 -/- fibroblasts and subjected to sequencing. IGV browser snapshot of RNA-seq, peaks indicate relative expression levels at SMARCC2 (A), DDB2 (B) and EPB41L2 (C) genes. EPB41L2 is shown as sashimi plot (Katz et al. 2015). Venn diagram representing the overlap between the change in expression to that of splicing in F1-II:1 -/- (D) and F2-II:1 -/- (E) and the gene-expression overlap to abnormal splicing overlap between the two probands (F).
